# Testing-on-a-probe biosensors reveal association of early SARS-CoV-2 total antibodies and surrogate neutralizing antibodies with mortality in COVID-19 patients

**DOI:** 10.1101/2020.11.19.20235044

**Authors:** He S. Yang, Sabrina E. Racine-Brzostek, Mohsen Karbaschi, Jim Yee, Alicia Dillard, Peter A.D. Steel, William T. Lee, Kathleen A. McDonough, Yuqing Qiu, Thomas J. Ketas, Erik Francomano, P. J. Klasse, Layla Hatem, Lars Westblade, Heng Wu, Haode Chen, Robert Zuk, Hong Tan, Roxanne C. Girardin, Alan P. Dupuis, Anne F. Payne, John P. Moore, Melissa M. Cushing, Amy Chadburn, Zhen Zhao

**Author notes:** Corresponding author: Zhen Zhao, PhD, 525 East 68^th^ Street; F-701; New York, NY 10065. **Abbreviation:** ACE2: angiotensin-converting enzyme 2; BSL: biosafety level; COVID-19: Coronavirus Disease-2019; Cy5-SA-PS: Cy5-Streptavidin-polysaccharide; C_*T*_: cycle threshold; DAED: days after Emergency Department presentation; DAOS: days after onset of symptoms; ED: Emergency Department; EUA: emergency use authorization; FDA: U.S. Food and Drug Administration; HR: hazard ratio; IC50: half maximal inhibitory concentration; IQR: interquartile range; NYP: New York Presbyterian; PRNT: plaque reduction neutralization test; PsV: pseudovirus neutralization test; RBD: receptor-binding domain; RFU: relative fluorescence unit; RT-PCR: real-time reverse transcription polymerase chain reaction; SARS-CoV-2: severe acute respiratory syndrome coronavirus 2; SD: standard deviation; SMCC: succinimidyl 4-[N-malemidomethyl]cyclohexan-1-carboxylate; SNAB: surrogate neutralizing antibody; SPDP: succinimydyl 6-[3-[2-pyridyldithio]-proprionamido] hexanoate; TAb: total antibody; TOP: Testing-on-a-probe; WCMC: Weill Cornell Medical Center.

## Abstract

The association of mortality with early humoral response to SARS-CoV-2 infection within the first few days after onset of symptoms (DAOS) has not been thoroughly investigated partly due to a lack of sufficiently sensitive antibody testing methods. Here we report two sensitive and automated testing-on-a-probe (TOP) biosensor assays for SARS-CoV-2 viral specific total antibodies (TAb) and surrogate neutralizing antibodies (SNAb), which are suitable for clinical use. The TOP assays employ an RBD-coated quartz probe using a Cy5-Streptavidin-polysacharide conjugate to improved sensitivity and minimize interference. Disposable cartridge containing pre-dispensed reagents requires no liquid manipulation or fluidics during testing. The TOP-TAb assay exhibited higher sensitivity in the 0-7 DAOS window than a widely used FDA-EUA assay. The rapid (18 min) and automated TOP-SNAb correlated well with two well-established SARS-CoV-2 virus neutralization tests. The clinical utility of the TOP assays was demonstrated by evaluating early antibody responses in 120 SARS-CoV-2 RT-PCR positive adult hospitalized patients. Higher baseline TAb and SNAb positivity rates and more robust antibody responses were seen in patients who survived COVID-19 than those who died in the hospital. Survival analysis using the Cox Proportional Hazards Model showed that patients who were TAb and SNAb negative at initial hospital presentation were at a higher risk of in-hospital mortality. Furthermore, TAb and SNAb levels at presentation were inversely associated with SARS-CoV-2 viral load based on concurrent RT-PCR testing. Overall, the sensitive and automated TAb and SNAb assays allow detection of early SARS-CoV-2 antibodies which associate with mortality.

## 1. Introduction

Ten months after the emergence of Coronavirus Disease 2019 (COVID-19) and the identification of more than 40 million confirmed cases (Dong et al. 2020), many clinical and biologic aspects of the disease are not yet understood (Callaway et al. 2020). It is unknown why the initial clinical presentation does not necessarily correlate with subsequent disease severity and why some patients, even those who are young or are without significant co-morbidities, die from COVID-19 whereas others recover. Analyses of risk factors for a poor outcome in hospitalized COVID-19 patients (Li et al. 2020; Ruan et al. 2020; Wolff et al. 2020; Zheng et al. 2020) have largely focused on patient clinical presentation, demographics, comorbidities, and non-specific laboratory testing results (Wiersinga et al. 2020). However, COVID-19 serologic assays that evaluate patient humoral response to the severe acute respiratory syndrome coronavirus 2 (SARS-CoV-2) are more disease specific and thus are likely to correlate better with disease severity and/or survival (Klasse and Moore 2020). While many studies have reported that the magnitude of antibody response to SARS-CoV-2 tracks with the severity of COVID-19, these differences are generally seen more than 2-3 weeks after the onset of symptoms (Long et al. 2020; Lynch KL 2020; Yang et al. 2020; Zhao et al. 2020). Variations in the early humoral immune response to SARS-CoV-2, which may be more predictive of clinical outcome, have not been thoroughly investigated, due at least in part, to the lack of serologic testing methods that are sufficiently sensitive during the key early days of infection (Tang et al. 2020; Yang et al. 2020). Moreover, few studies have focused on the association of early antibody response and patient outcome, including mortality (Atyeo et al. 2020).

Most neutralizing antibodies (NAbs) against SARS-CoV-2 block the receptor-binding domain (RBD) of the spike (S) glycoprotein from interacting with human angiotensin-converting enzyme 2 (ACE2), thereby preventing virus entry the host cell (Ju et al. 2020). The presence of NAbs early after infection may be important in avoiding severe disease manifestations by limiting the number of host cells that become productively infected. However, measuring SARS-CoV-2 NAb activity during the acute phase of COVID-19 is difficult for clinical laboratories; the assays often require specialized equipment and are labor intensive, time consuming (4 days) procedures, and hence throughput is low. For instance, the conventional virus neutralization test requires biosafety level (BSL)-3 containment facilities for handling live virus and takes several days to complete (Lee WT 2020). Although the pseudovirus neutralization test and ELISA-based surrogate neutralization assays can be performed in the more widely available BSL-2 laboratories, they are still labor intensive and time consuming (Schmidt et al. 2020), and hence problematic for generating real time results in the clinical arena.

In this study, we developed two rapid, sensitive and fully automated testing-on-a-probe (TOP) assays to detect and quantify: i) total SARS-CoV-2 antibodies (TAb); and, ii) surrogate neutralizing antibodies (SNAb) that inhibit RBD-ACE2 interactions. The TOP assay employs a receptor-binding domain (RBD)-coated quartz tip biosensor with Cy5-Streptavidin-polysaccharide (Cy5-SA-PS) conjugate, which facilitates the capture of target molecules, minimizes the effect of any interfering substances in the serum and enhances the signal intensity. We then performed analytical and clinical validation of the TOP assays. The clinical utility of these novel assays was further investigated to explore whether the timing and magnitude of the SARS-CoV-2 antibody response are associated with COVID-19 mortality.

## 2. Experimental methods

### 2.1. Cy5-Streptavidin-high molecular weight polysaccharide (Cy5-SA-PS) conjugate

The sensitivity of the new assays is enhanced by improving the conjugation chemistry to increase the binding signal and minimizing the non-specific binding. Details are described in the **Supplemental Materials**.

### 2.2. Testing-on-a-probe (TOP) SARS-CoV-2 total RBD antibody assay (TAb)

The TOP-TAb assay measures the overall binding between SARS-CoV-2 antibodies and the receptor-binding domain (RBD) of the virus spike (S) protein (**Figure 1A**). The assay is comprised of RBD pre-coated probes and preloaded reagent strips. To construct the biosensor probe, silane treated hydrophobic glass pins were coated with recombinant SARS-CoV-2 RBD protein (Sino Biological, Beijing, China). Biosensor probes were then exposed to PBS with 15% sucrose, as a preservative, and dried in an oven for 30 minutes at 40°. Strips were constructed of biotinylated RBD using EZ-Link NHS-LC-LC-Biotin (Thermo Fisher Scientific, Waltham, MA, USA) and Cy5-SA-PS conjugate. Finally, probes and strips were assembled together as a single cartridge (ET Healthcare, Palo Alto, CA).

**Figure 1.**
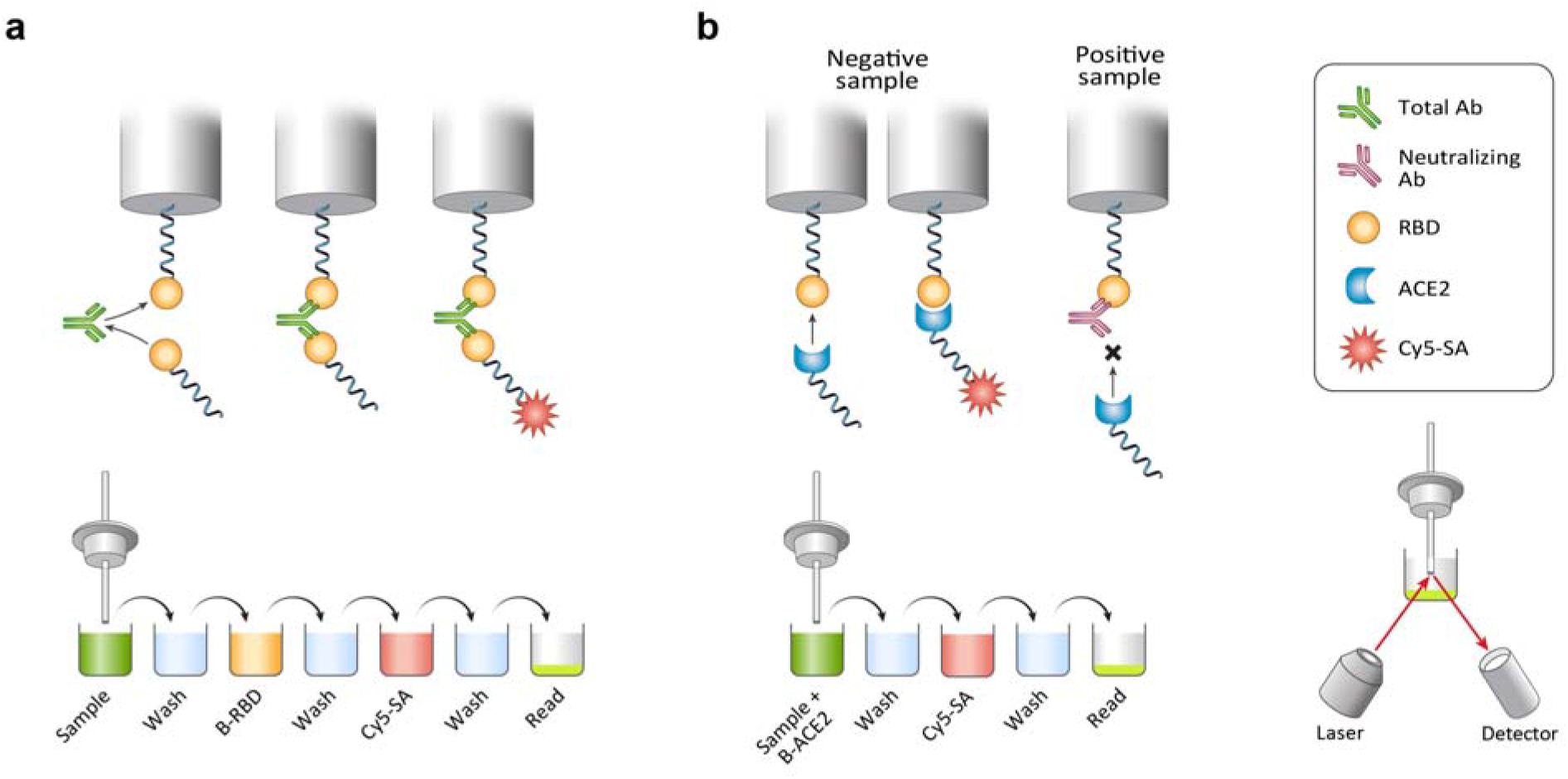
Principles of fully automated, testing-on-a-probe (TOP) Total Antibody (TAb) and Surrogate Neutralizing Antibody (SNAb) Assays. The assay cartridge consists of an RBD pre-coated probe and preloaded reagent microwells. (A) TAb assay: The instrument sequentially transfers and incubates RBD pre-coated biosensor probe in a well with diluted sample to capture SARS-CoV-2 specific antibodies, a wash well, a biotinylated RBD well, a wash well, a Cy5-Streptavidin-polysacharide (Cy5-SA-PS) well, and a wash well. At the end, the probe is transferred to a well where the fluorescence bound on the tip of the biosensor is measured. (B) SNAb assay: the instrument sequentially transfers and incubates RBD pre-coated biosensor probe in a well containing a mixture of patient sample and biotinylated ACE2, a wash well, a Cy5-SA-PS well and a wash well. At the end, the biosensor probe is transferred to a read well where the fluorescence bound on the biosensor tip is measured. (C and D) Assay sensitivity enhancement by conjugation of Cy5-SA to a high molecular weight PS. Samples of SARS-CoV-2 negative human serum spiked with monoclonal SARS-CoV-2 IgG (C) or IgM (D) at different concentrations were measured on the TOP-TAb assay with Cy5-SA or Cy5-SA-PS as the signaling element. The Cy5-SA-PS showed enhanced signal sensitive by up to 20-fold and reduced background noise by 3-fold compared to Cy5-SA.

The TAb assay is fully automated on the Pylon 3D analyzer (ET Healthcare). The instrument’s pipetting system dilutes the patient sample (26 µL) in PBS (1:5) and dispenses the diluted the solution into a microwell. The instrument probe is moved through sequential incubations in a series of wells containing diluted patient sample, biotinylated RBD and finally the Cy5-SA-PS conjugate. After each incubation, the instrument carries out a brief wash sequence. The instrument detects the fluorescence of the B-RBD/Cy5-SA bound on the probe tip using a built-in optical system as the last step of the assay. The total assay time is 16 min.

### 2.3. TOP SARS-CoV-2 surrogate neutralizing antibody assay (SNAb)

Designed as a competitive binding assay, the TOP-SNAb (**Figure 1B**), is based on the anti-SARS-CoV-2 antibody-mediated inhibition of the interaction between the ACE2 receptor protein and the RBD. The SNAb assay is also fully automated and can be performed using the same instrument as TAb. The assay readout is the percentage of RBD-ACE2 binding, which inversely correlates with the SNAb binding inhibition (neutralizing activity) as described by another SNAb assay (Tan et al. 2020). The percentage of RBD-ACE2 binding is defined as %B/B0 = (sample RFU/negative control RFU) *100%. The SNAb instrument pipette mixes the patient sample (26 µL) with a biotinylated ACE2 reagent (SinoBiological, 10108-H08H-B) in a well, after which a RBD pre-coated biosensor probe (as described for the TAb assay) is transferred to the well containing the patient sample and biotinylated ACE2 mixture. Following a 12 minute incubation, the instrument transfers the RBD probe to the well containing Cy5-SA-PS. After each incubation, the instrument carries out a brief wash sequence of 15 seconds. The instrument then detects the fluorescence of biotinylated ACE2/Cy5-SA bound on the probe tip and reports it as previously described. The total assay time is 18 min.

### 2.4. The Roche Elecsys Anti-SARS-CoV-2 assay

The Roche Elecsys Anti-SARS-CoV-2 assay (Roche TAb; Roche Diagnostics, Indianapolis, IN) was performed on the Roche Cobas e411 (Roche Diagnostics). This assay received Emergency Use Authorization (EUA) approval from the United States Food and Drug Administration (FDA) and was used for comparison with the TOP-TAb assay.

### 2.5. The plaque reduction neutralization test (PRNT) and pseudo virus neutralization test (PsV)

The PRNT assay detects viral specific antibodies based on their ability to neutralize their cognate viral infections in Vero E6 cells (C1008, ATCC CRL-1586)(Lee WT 2020). The PsV assay detects neutralizing antibodies based on their ability to inhibit the entry of SARS-CoV-2 pseudovirus into 293T/ACE2cl.22 cells (obtained from Paul Bieniasz, The Rockefeller University, New York) (Schmidt et al. 2020). The PsV assay was performed as described previously (Schmidt et al. 2020) with minor modifications. The titers of PRNT and PsV were reported as PRNT 50 and the half maximal inhibitory concentration (IC50), respectively. The details of these two NAb assays are described in the **Supplementary Materials**.

### 2.6. SARS-CoV-2 RT-PCR assay and the viral load quantification

SARS-CoV-2 RT-PCR testing was performed as part of routine patient care at the time of the patient’s emergency department (ED) visit on the Cobas 6800 RT-PCR system (Roche Molecular Systems, Inc., Branchburg, NJ), an assay which received EUA approval from the FDA. The SARS-CoV-2-specific target, ORF1ab, is amplified in this assay and the C_*T*_ values were obtained for each patient’s RT-PCR result. The C_*T*_ value represents the number of replication cycles required for sufficient gene amplification to produce a fluorescent signal that crosses a predefined threshold. Analysis of C_*T*_ values has been previously described (Magleby et al. 2020a). For analysis purposes, patients were categorized into one of three cohorts, based on quantitative C_*T*_ values: high viral load (C_*T*_ <25), medium viral load (C_*T*_ 25-30), and low viral load (C_*T*_ >30) based on the previous analysis from our institution (Magleby et al. 2020a).

### 2.7. Sources of serum specimens and data acquisition

The clinical study was approved by the Institutional Review Board (#20-03021671) of Weill Cornell Medicine (WCM) with a waiver of informed consent. Neutralizing antibody testing at the Wadsworth Center was done under a declared Public Health Emergency with a waiver from the NYSDOH Institutional Review Board.

The retrospective clinical study included a primary study cohort of 120 adult patients who presented to the ED and were subsequently hospitalized at NewYork Presbyterian/Weill Cornell Medical Center (NYP/WCMC) from March 8 to April 7, 2020 (**Table 1**). All patients had a nasopharyngeal swab sample that tested positive by SARS-CoV-2 real-time reverse transcription polymerase chain reaction (RT-PCR) within one day of their ED visit. All 120 patients had a serum sample collected within one day of the initial hospital presentation for SARS-CoV-2 antibody analysis. Clinical data collected from the electronic medical record (Allscripts, Chicago, IL) included demographic information, estimated date of symptom onset, presenting symptoms (fever, cough, sore throat, shortness of breath, chest pain, diarrhea, headache, nausea, vomiting, body or muscle aches, fatigue/weakness, ageusia, anosmia, abdominal pain, rhinitis), comorbidities (cancer, obesity, type 2 diabetes, hypertension, hyperlipidemia, and chronic cardiac diseases), compromised immune status (post-transplant, chemotherapy, radiation and chronic corticosteroid use), SARS-CoV-2 RT-PCR results, intubation status and in-hospital mortality. The last day of study follow-up by chart review was September 6, 2020.

**Table 1.** Baseline characteristics of hospitalized patients stratified by death.

| **Variable** | **Categories/Statistics** | **Death**<br>**(n=32)** | **Survival**<br>**(n=88)** | **P-value** |
| --- | --- | --- | --- | --- |
| Intubation | n. (%) | 20 (62.5%) | 18 (20.5%) | <0.001<sup>C</sup> |
| Gender | F | 7 (21.9%) | 35 (39.8%) | 0.069<sup>C</sup> |
|  | M | 25 (78.1%) | 53 (60.2%) |  |
| Age | Mean (SD) | 76.6 (12.5) | 61.8 (14.9) | <0.001<sup>T</sup> |
|  | Median (IQR) | 78.5 (70, 86.2) | 62.5 (53, 71) |  |
|  | Range | (46, 96) | (24, 92) |  |
| Race | Asian | 4 (12.5%) | 3 (3.4%) | 0.082<sup>F</sup> |
|  | Black/AA | 2 (6.2%) | 13 (14.8%) |  |
|  | Declined | 6 (18.8%) | 20 (22.7%) |  |
|  | Other | 2 (6.2%) | 18 (20.5%) |  |
|  | Unknown | 3 (9.4%) | 8 (9.1%) |  |
|  | White | 15 (46.9%) | 26 (29.5%) |  |
| DAOS at initial hospital presentation | Mean (SD) | 7.8 (5.3) | 7.5 (4.6) | 0.98<sup>E</sup> |
|  | Median (IQR) | 7 (4, 14) | 7 (4, 10) |  |
|  | Range | (0, 21) | (1, 21) |  |
| Cancer | n. (%) | 12 (37.5%) | 9 (10.2%) | <0.001<sup>C</sup> |
| Other Comorbidities<sup>*</sup> | n. (%) | 26 (81.2%) | 61 (69.3%) | 0.20<sup>C</sup> |
| Compromised immune status<sup>#</sup> | n. (%) | 6 (18.8%) | 8 (9.1%) | 0.20<sup>F</sup> |
| Abdominal pain | n. (%) | 2 (6.2%) | 6 (6.8%) | >0.99<sup>F</sup> |
| Fever | n. (%) | 17 (53.1%) | 60 (68.2%) | 0.13<sup>C</sup> |
| Chest pain | n. (%) | 1 (3.1%) | 1 (1.1%) | 0.46<sup>F</sup> |
| Shortness of breath | n. (%) | 23 (71.9%) | 48 (54.5%) | 0.088<sup>C</sup> |
| Cough | n. (%) | 14 (43.8%) | 45 (51.1%) | 0.47<sup>C</sup> |
| Diarrhea | n. (%) | 2 (6.2%) | 15 (17%) | 0.23<sup>F</sup> |
| Headache | n. (%) | 1 (3.1%) | 5 (5.7%) | >0.99<sup>F</sup> |
| Rhinitis | n. (%) | 0 (0%) | 2 (2.3%) | >0.99<sup>F</sup> |
| Nausea/Vomiting | n. (%) | 1 (3.1%) | 14 (15.9%) | 0.068<sup>F</sup> |
| Ageusia or Anosmia | n. (%) | 1 (3.1%) | 3 (3.4%) | >0.99<sup>F</sup> |
| Body or muscle aches | n. (%) | 2 (6.2%) | 16 (18.2%) | 0.15<sup>F</sup> |
| Fatigue/Weakness | n. (%) | 9 (28.1%) | 25 (28.4%) | 0.98<sup>C</sup> |
| Sore throat | n. (%) | 1 (3.1%) | 1 (1.1%) | 0.46<sup>F</sup> |

Test: C: Chi-square; E: Exact Wilcox; F: Fisher; T: T.test
\*Other comorbidities included hypertension, type 2 diabetes, coronary artery disease, hyperlipidemia, and obesity;

## 3. Results

### 3.1. Analytical validation of the TOP-TAb and TOP-SNAb assay

The principles of the fully automated biosensor TOP-TAb and TOP-SNAb assays are illustrated in **Figure 1A and B**. Cy5-SA-PS showed enhanced sensitivity and reduced background. The Cy5-SA-PS generated up to 20- and 16-times higher fluorescence signal from monoclonal SARS-CoV-2 IgG and IgM, respectively, compared to Cy5-SA. The background fluorescence signal of Cy5-SA-PS was 3-times lower than that of Cy5-SA (**Figure 1C and D**). The cutoff values of both methods were determined by the mean plus 6 standard deviation (SD) using pre-COVID-19 samples collected in July 2019 (n = 50 for TAb and n = 20 for SNAb). The cutoff values of TAb and SNAb were 20 RFU and 85% B/B0, respectively. Additional pre-COVID 19 serum samples (n = 163 for TAb and n = 27 for SNAb) collected in July 2019 were tested to validate the specificity, which was 99.4% and 100% for TAb and SNAb, respectively. The imprecision was determined by running the high and low levels of quality control samples 2-5 times per day on 3-5 different days. The imprecision of TAb and SNAb measured by coefficient of variation (CV) was 8% and 12%, respectively. The linearity was performed by serial dilution of high TAb and SNAb samples and at least 5 dilution points were used. Linearity was analyzed by simple linear regression. TAb and SNAb had linearity of 8-7494 RFU (R^2^ = 0.98) and 3-96% B/B0 (R^2^ = 0.99), respectively (**Supplemental Figure 1**).

### 3.2. Correlations of the TOP-SNAb assay with two well-established SARS-CoV-2 neutralizing antibody assays

To test the accuracy of the TOP-SNAb assay, we compared SNAb with two other established NAb assays, the plaque reduction neutralization test (PRNT) and pseudovirus neutralization test (PsV), using 46 remnant samples randomly collected from COVID-19 convalescent outpatients. Data generated in the latter two assays were concordant, with a correlation coefficient (r) of 0.77 (p < 0.0001; **Figure 2A**). The percentage of ACE2-RBD binding in the SNAb assay, which is an inverse surrogate for virus-neutralization (SNAb binding inhibition), was then shown to correlate well with both the PRNT (r = −0.82, p < 0.0001) and PsV assays (r = −0.80, p < 0.001) (**Figure 2B and C**).

**Figure 2.**
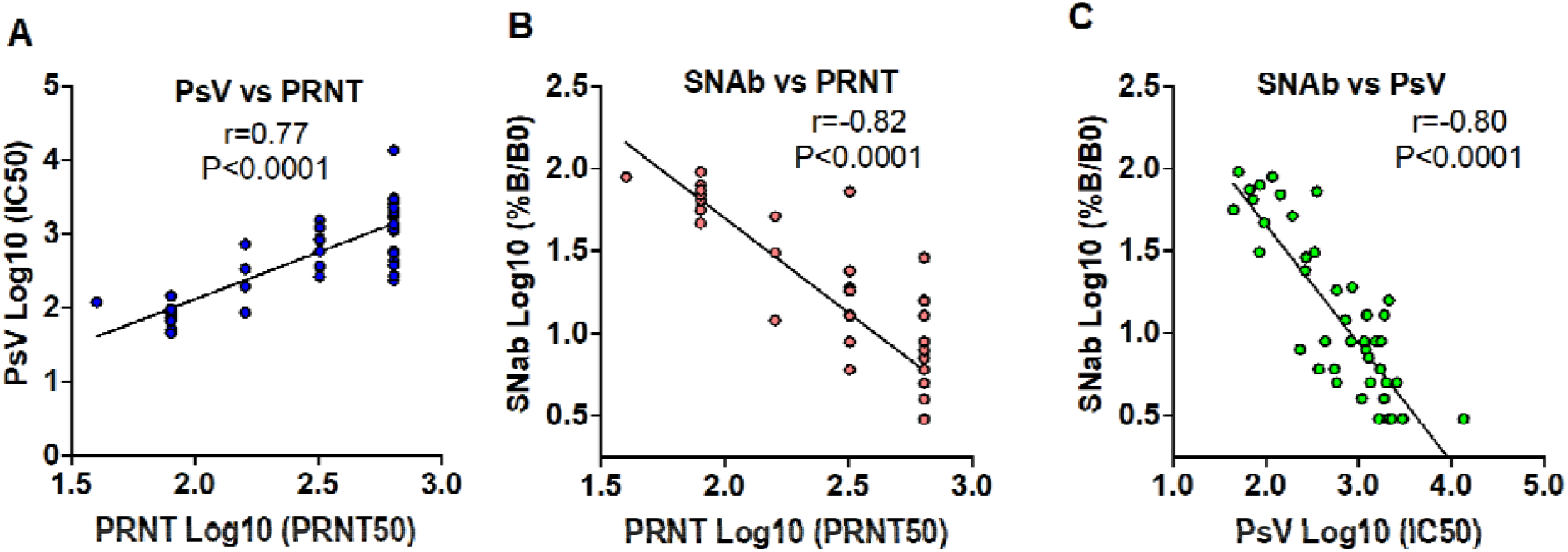
Correlation of the TOP-SNAb assay with two well-established SARS-CoV-2 NAb assays. The correlations between the PsV and PRNT, between TOP-SNAb and PRNT, and between TOP-SNAb and PsV are shown in Figure 2A, B, C, respectively. The readout of TOP-SNAb is the percentage of RBD-ACE2 binding, which inversely correlates with the SNAb binding inhibition (neutralizing activity). The titers of PRNT and PsV were reported as PRNT 50 and IC50, respectively. The results were presented as Log10 scale. Correlations between two assays were assessed by Spearman correlation coefficient.

### 3.3. Validation of clinical sensitivity of TOP-TAb and TOP-SNAb assays

As an evaluation of clinical sensitivity, 116 serum samples from 39 patients were analyzed to compare TOP-TAb and TOP-SNAb with a widely used FDA-approved (EUA) total antibody assay (Roche TAb). The detection rates of the three assays during different periods: days after onset of symptoms (DAOS) and days after initial ED presentation (DAED) are shown in **Figure 3A and Supplementary Table 1 and 2**. On the day of initial presentation, the TOP-TAb assay [61.5% (95% CI: 45.9-75.1%)] was significantly more sensitive than the Roche TAb assay [33.3% (95% CI: 20.6-49.1%), p = 0.023]. The TOP-SNAb assay was also more sensitive than Roche TAb, but not to a significant extent (p = 0.100). When used in the earliest stage of infection (0-7 DAOS), the sensitivity of TOP-TAb [57.1% (95% CI: 40.8-72.0%)] was still significantly higher than that of the Roche TAb [26.5% (95% CI: 14.4-43.3), p = 0.015].

**Figure 3.**
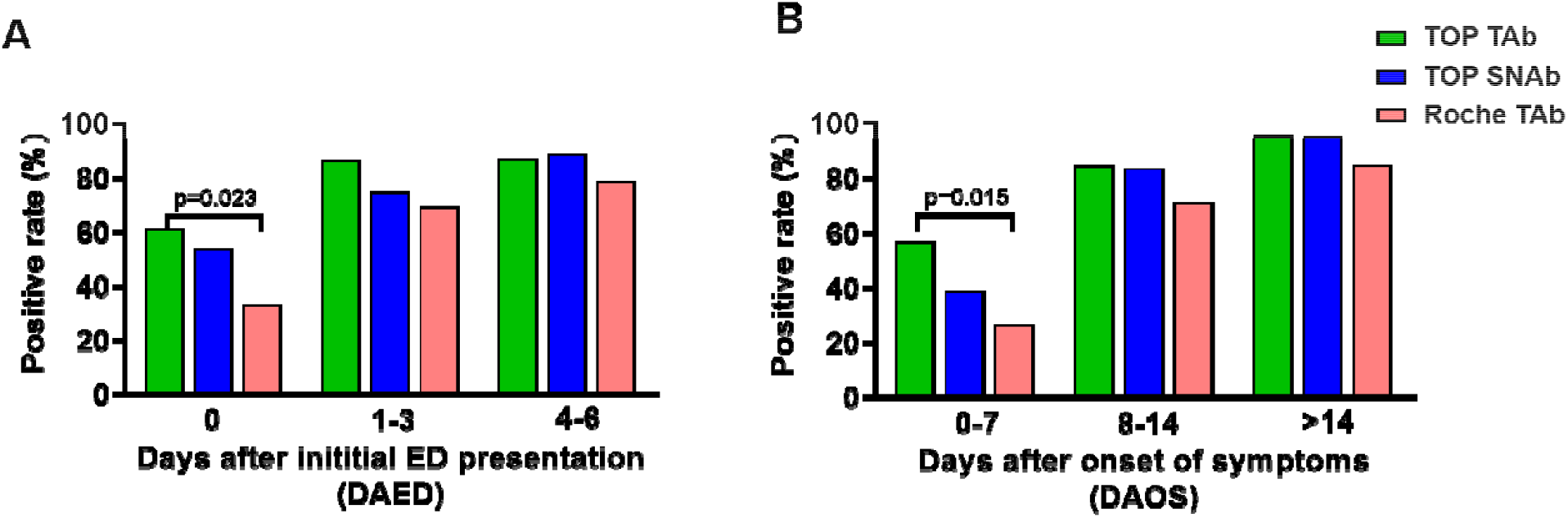
Detection rates of SARS-CoV-2 TOP-TAb, TOP-SNAb, and Roche Tab. Detection rates were evaluated based on (A) days after initial ED visit (DAED) and days after onset of symptoms (DAOS).

As expected, the sensitivity of each antibody assay increased substantially at later testing times (> 3 DAED and >7 DAOS); there were no significant differences between the three assays during this later period (**Figure 3B and Supplementary Table 1 and 2**). The most prominent advantage of the TOP-TAb and TOP-SNAb assays is their superior ability to detect antibodies in the critical early window from 0-7 DAOS, when patients first present for clinical care.

### 3.4. Evaluation of clinical utility: Association of TAb levels and SNAb binding inhibition at initial hospital ED presentation with in-hospital mortality

To further evaluate the clinical utility of the more sensitive and automated assays within the first few days of infection, TAb and SNAb were measured in SARS-CoV-2 RT-PCR positive patients who had the ED day 0 serum sample collected during their initial hospital presentation. Of the 120 patients, 31.7% (38/120) were intubated and 26.7% (32/120) died during hospitalization. The median DAOS at the time of hospital presentation was 7 days (IQR: 4-10). Demographic, clinical presentation and comorbidity data are shown in **Table 1 (**stratified by survival status) and **Supplementary Table 3** (stratified by intubation status). Intubation, age and cancer were significantly higher in patients who died than the survivors (p < 0.001, Table 1). SNAb data were missing in 6 samples due to an insufficient quantity of serum.

Patient antibody status in the TAb assay was categorized into positive (signal > 20 IFU, the assay cutoff) and negative (signal ≤ 20 IFU) groups, and similarly for the SNAb assay (positive = %B/B0 ≤ 85%, the assay cutoff; negative = %B/B0 > 85%, see methods section). The TAb positivity rate was significantly higher for the patients who survived (63.6%; 56/88) than for patients who died (34.4%, 11/32, p = 0.007, **Figure 4A**). Similarly, the SNAb positivity rate for patients who survived (61.2%, 52/85) was also higher than for those who died (24.1%, 7/29, p = 0.001, **Figure 4A**). Furthermore, the TAb levels and SNAb binding inhibition were significantly higher in the patients who survived than who died (p = 0.004 and p = 0.004, respectively, **Figure 4B, 4C**). In contrast, no significant differences in antibody positivity rates or levels of TAb and SNAb in the initial ED samples were observed when comparing intubation status. However, the SNAb positivity rate was lower for patients who were intubated than for those who were not. (p=0.026, **Supplementary Figure 2**).

**Figure 4.**
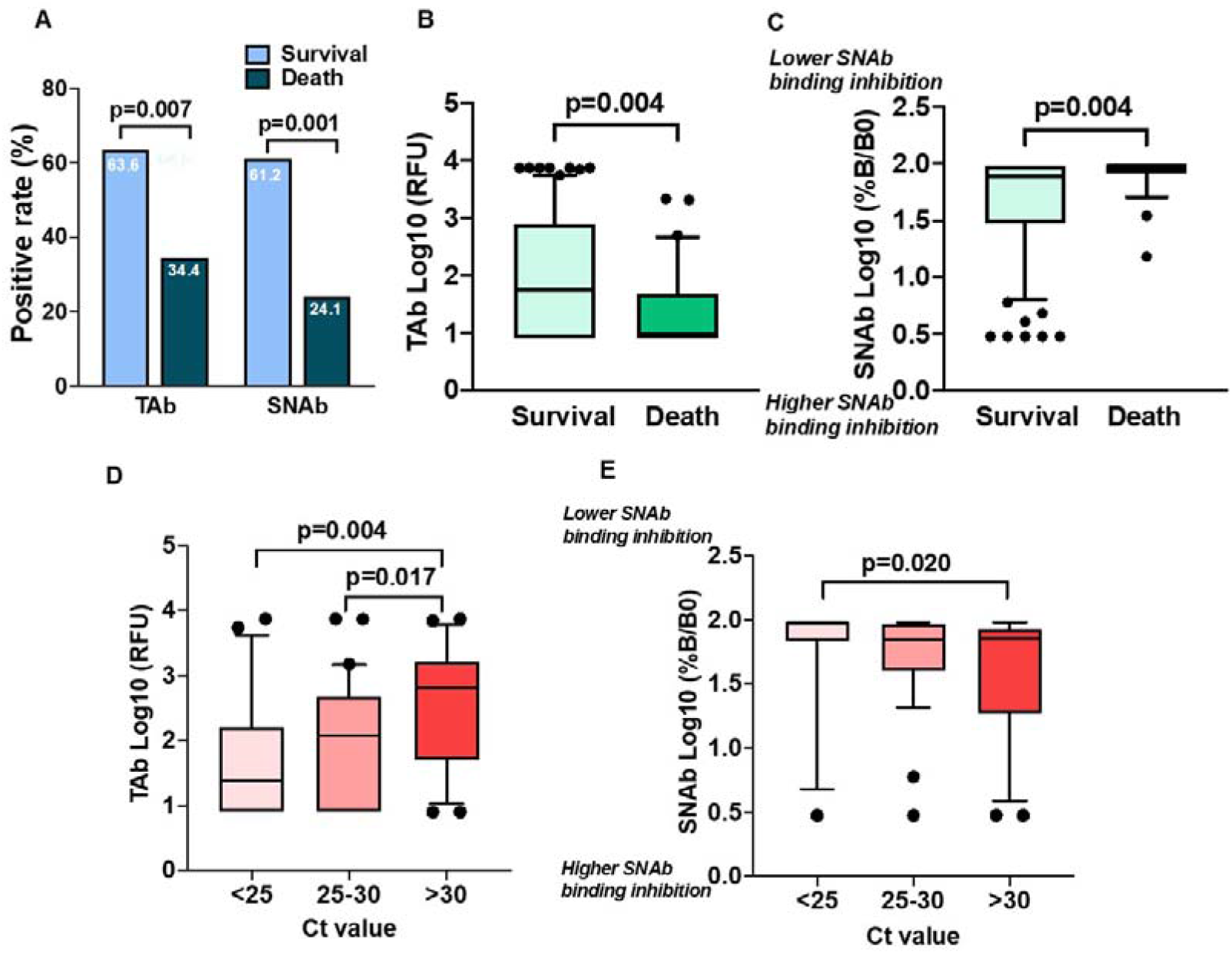
TOP-TAb and TOP-SNAb at initial hospital ED presentation stratified by in-hospital mortality and viral load. (A) At the initial hospital ED presentation, TAb and SNAb positive rates were higher in patients who survived than who died. (B) TAb level and (C) SNAb binding inhibition were higher in patients who survived than who died. Association of (D) TAb and (E) SNAb with SARS-Cov-2 ORF1ab target C_*T*_ values at the time of hospital ED presentation. Data were expressed as Log10 scale with box and whisker (10-90 percentile) plots.

Survival analysis was performed using the Cox Proportional Hazards Model adjusted for age and cancer comorbidity, which were two variables showing significant differences between death and survival groups in the univariate analysis (**Table 1**). Patients with negative TAb at the time of initial hospital presentation had a higher hazard of in-hospital mortality than those with a positive TAb [hazard ratio (HR) = 2.33 (95% CI: 1.09-5.00, p=0.029), **Figure 5A**]; a similar finding was made when the SNAb data were analyzed in the same way [HR = 3.25 (95% CI: 1.33-7.94; p=0.01), **Figure 5B**].

**Figure 5.**
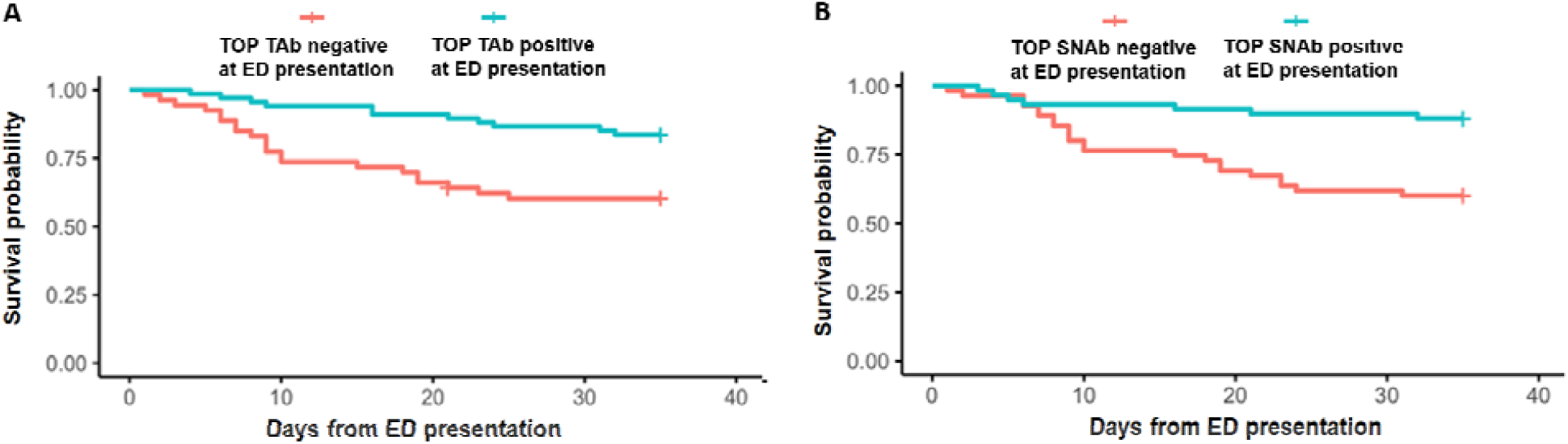
Survival probability among SARS-CoV-2 infected patients with positive and negative (A) TOP-TAb and (B) TOP-SNAb at initial hospital ED presentation. Data were analyzed using Cox proportional hazards regression adjusting for age and cancer comorbidity.

### 3.5. Higher TAb levels and SNAb binding inhibition were associated with lower viral loads at ED presentation

To further validate whether TAb and SNAb levels associated with SARS-CoV-2 viral load, the initial nasopharyngeal swab SARS-CoV-2 RT-PCR C_*T*_ values determined using the Cobas SARS-CoV-2 assay were obtained from 73 of 120 (60.8%) patients. All C_*T*_ values generated using other RT-PCR assay systems were excluded for consistency. Using a method described elsewhere, the resulting data sets were grouped as follows: high viral load (C_*T*_ value < 25, n = 21, 28.8%), medium viral load (C_*T*_ value 25-30, n = 31, 42.5%), low viral load (C_*T*_ value > 30, n = 21, 28.8%). TAb (n= 73) and SNAb (n = 68) in samples collected within one day of the RT-PCR testing were compared with the C_*T*_ values. The analysis showed that the patients with low viral loads (C_*T*_ > 30) had significantly higher TAb levels than those with the medium (p = 0.017) and low viral loads groups (p=0.004, **Figure 4D**), respectively. Similarly, patients with low viral loads (C_*T*_ > 30) also had higher SNAb binding inhibition than the low viral load group (p = 0.020, **Figure 4E**).

## 4. Discussion

In this study, we report that novel TOP-TAb and TOP-SNAb assays can detect SARS-CoV-2 viral specific antibodies early after the onset of symptoms, notably more sensitive than a commonly used SARS-CoV-2 serology assays. The greater sensitivity of these new assays allows for the detection of SARS-CoV-2 antibody responses on the first day of hospital visit that associate with COVID-19 mortality. In this initial care context, the rapidity, simplicity and sensitivity of the TOP assays are all advantages over previous methods. Both TAb and SNAb assays yield results in 16-18 minutes and the instrument is easy to operate in a routine clinical laboratory without the need for BSL-3 containment. The new SNAb assay offers a great opportunity to incorporate the measurement of SNAb into a clinical assessment of patient’s risk while the patient is waiting in the ED or a physician’s office.

The TOP biosensor is a versatile sensing method. Unlike most of other biosensor technologies that transport samples to sensors, the TOP method brings the sensor to a sample to achieve high sensitivity and high precision. Specifically, to test SARS-CoV-2 TAb and SNAb, the TOP assays utilize an RBD-coated quartz tip that moves through microwells on an unitized disposable test strip with pre-dispensed reagents. No reagents are dispensed during the assay procedure eliminating the need for fluid handling subsystems in the instrument. Whole blood, serum and plasma specimens can all be tested using TOP biosensors. The small surface area of the pre-coated tip (1 mm diameter) allows for small sample volume (26 µL), low reagent consumption and, more importantly, negligible interaction with interference from patient samples. The fluorescence signal is enhanced by a Cy5-SA-PS conjugate. The high-molecular-weight PS serves as an inert carrier of multiple Cy5-SA molecules (∼ 30 SAs / PS) and amplifies the signal by approximately 20-fold compared to unconjugated Cy5-SA. Background signal is negligible since the PS is hydrophilic and has minimal interaction with the tip surface. Utilization of TOP with the Cy5-SA-PS conjugate results in an improved signal-to-noise ratio, i.e. high sensitivity, for the detection of SARS-CoV-2 antibodies. Although the TOP assays in this study were not designed to differentiate antibody isotypes and are sensitive to only SARS-CoV-2 RBD or spike protein specific antibodies, the approach described in this study simplifies assays and their development process significantly. New assay development can be readily accomplished by using the same biosensor probe designed to generate signal in response to a specific SARS-CoV-2 antibody isotype and target other SARS-CoV-2 specific antigens.

Utilizing the TOP assays, we demonstrate, for the first time, that SARS-CoV-2 TAb and SNAb upon initial presentation are risk indicators for in-hospital mortality, and higher antibody levels are associated with a lower viral load. There has been significant controversy over the antibody response in patients with different levels of COVID-19 disease severity and different outcomes (Klasse and Moore 2020). Multiple studies have shown that SARS-CoV-2 specific antibody levels, such as IgM, IgG or NAb are elevated in patients with a more severe disease courses (Lynch KL 2020; Wang et al. 2020a; Wang et al. 2020b; Zhou et al. 2020; Zohar and Alter 2020). However, a few other publications have reported no significant differences in SARS-CoV-2 antibody responses between severe and non-severe patients (Phipps et al. 2020; Ren et al. 2020). These discrepancies could involve a variety of factors, including the patient populations, the definitions of disease severity, the serology assays used, the isotypes of antibody or target epitopes being detected, and the timing in the disease course when the antibody testing is performed. Here, we focused on the association, using the sensitive TOP assays, between Tab and SNAb on the first day of hospital visit and subsequent mortality. Our findings are congruent with recent reports that early immunologic protection in COVID-19 patients is important. For instance, Atyeo et al. identified that an early SARS-CoV-2-specific humoral signature profile, including levels of S-specific IgM and IgA, N-specific complement activity, IgM and IgA1, is associated with later disease outcome (Atyeo et al. 2020). Thus, it is reasonable to postulate that if SARS-CoV-2 TAbs and NAbs appear early in the disease process, they may play a protective role directly or indirectly suppressing SARS-CoV-2 replication. Antibody response kinetics may differ in patients who survive versus those who die from COVID-19. For example, patients with low or undetectable levels of SARS-CoV-2 antibodies at an early stage may not be able to suppress the initial phase of virus replication; the resulting seeding of multiple foci of infection could then drive the emergence of high viral loads, which could in turn trigger the production of a much stronger later antibody response. However, these high-titer antibodies may appear too late to prevent severe disease and death. These hypotheses are concordant with our observation that patients with relatively high antibody levels early in the disease progress also had lower SARS-CoV-2 viral loads, particularly as viral load is independently associated with in-hospital mortality (Magleby et al. 2020b; Pujadas et al. 2020; Westblade LF 2020).

The associations between an early antibody response to SARS-CoV-2, initial viral load and eventual in-hospital survival are consistent with strong, early humoral immunity countering SARS-CoV-2 replication in a way that benefits disease control. A recent study of 35,000 hospitalized patients with COVID-19 showed that early transfusion of convalescent plasma (within 3 days of diagnosis) with high antibody levels was associated with reduced mortality (Joyner MJ 2020). The lack of an association with intubation may reflect the variability and evolution in our intubation practice during the height of the pandemic in New York City (Tobin M.J. 2020). These findings further argue for the clinical benefits of suppressing SARS-CoV-2 replication via specific antibodies very early during the disease course (i.e., the first week post onset of symptoms). The early identification of high-risk patients, (i.e., those who do not generate early and strong antibody responses of their own) could have significant implications for managing their immediate and future care.

## 5. Conclusion

We report a novel, rapid, highly sensitive and fully automated biosensor technology (TOP) that is easily adaptable to the clinical laboratory setting. These new assays allow the detection of early SARS-CoV-2 antibodies on the first day of a hospital or clinic visit and show that the levels of SARS-CoV-2 total and functional antibodies are inversely associated with subsequent COVID-19 mortality. Application of our new assays has led to the discovery of new information on the acute humoral immune response to SARS-CoV-2 infection and may have valuable prognostic and treatment implications relevant to patient care. Future studies will explore if these new sensitive and specific assays could potentially monitor the efficacy of antiviral therapies as well as assess antibody responses during vaccine trials.

## Data Availability

The data are available upon request

## Acknowledgement

This research was partially funded by a COVID-19 research grant from Weill Cornell Medicine. PJK, TJK, EF and JPM were supported by NIH grant R01 AI36082. We want to thank Hanna Rennert, Arryn R. Craney, Priya Velu for their effort on RT-PCR method development.

## Notes

**Conflict of interest statement:** ZZ received seed instruments and sponsored travel from ET HealthCare. ZZ received consulting fee from Roche Diagnostics. PAS received consulting fee from ET HealthCare.

### Competing Interest Statement

The authors have declared no competing interest.

