## Supplemental material for "Testing-on-a-probe biosensors reveal association of early SARS-CoV-2 total antibodies and surrogate neutralizing antibodies with mortality in COVID-19 patients"

He S. Yang^1,2^, Sabrina E. Racine-Brzostek^1,2^, Mohsen Karbaschi^3^, Jim Yee^2^, Alicia Dillard^1,2^, Peter A.D. Steel^2,4^, [William T. Lee](https://www.ncbi.nlm.nih.gov/pubmed/?term=Lee%20WT%5BAuthor%5D&cauthor=true&cauthor_uid=32505774)^5^, Kathleen A. McDonough^5^, Yuqing Qiu^6^, Thomas J. Ketas^7^, Erik Francomano^7^, P. J. Klasse^7^, Layla Hatem^1,2^, Lars Westblade^1^, Heng Wu^3^, Haode Chen^3^, Robert Zuk^3^, Hong Tan^3^, Roxanne C. Girardin^5^, Alan P. Dupuis II^5^, Anne F. Payne^5^, John P. Moore^7^, Melissa M. Cushing^1,2^, Amy Chadburn^1,2^, Zhen Zhao^1,2*^

^1^Department of Pathology and Laboratory Medicine, Weill Cornell Medicine, New York, NY, USA

^2^NewYork-Presbyterian Hospital/Weill Cornell Medical Campus, New York, NY, US

^3^ET HealthCare, Palo Alto, CA, USA

^4^Department of Emergency Medicine, Weill Cornell Medical Center, New York, NY, USA

^5^Diagnostic Immunology Laboratory, Wadsworth Center, New York State Department of Health, Albany, NY, USA

^6^Department of Population Health Sciences, Weill Cornell Medicine, New York, NY, USA

^7^Department of Microbiology and Immunology, Weill Cornell Medicine, New York, NY, USA

*Corresponding author:

Zhen Zhao, PhD

525 East 68^th^ Street; F-701; New York, NY 10065

**Supplementary material**

1. **Experimental methods**
   1.1. **Cy5-Streptavidin-high molecular weight polysaccharide (Cy5-SA-PS) conjugate**

For Cy5-SA-PS, a synthetic PS with a molecular weight of approximately 5 million Daltons with 5% of the sugar resides modified to contain amino groups was reacted with SPDP (succinimydyl 6-[3-[2-pyridyldithio]-proprionamido] hexanoate) (Invitrogen, CA, USA) in phosphate buffered saline (PBS; pH 7.4) for 1 hour, then purified with a PD-10 column (GE Healthcare, IL, USA). The thiols on the polysaccharide-SPDP were de-protected by adding dithiothreitol (Sigma Aldrich, MO, USA), incubating for 1 hour at room temperature, followed by purification of the polysaccharide-SH on a PD-10 column. Cy5-NHS (GE Healthcare) was linked to a streptavidin (Scripps Labs, CA, USA) by a standard method to contain 2 Cy5 dyes per streptavidin. Succinimidyl 4-[N-malemidomethyl]cyclohexan-1-carboxylate (SMCC; Thermo Fisher Scientific) was then incubated with the streptavidin at a 10 to 1 molar ratio for 1 hour at room temperature in PBS, followed by purification on a PD-10 column. The polysaccharide-SH and Cy5-Streptavidin-SMCC were mixed at a 1 to 1 weight ratio and allowed to react overnight at room temperature. N-ethylmaleimide (Sigma Aldrich) was then added to react with residual thiols and after 30 minutes the conjugate was purified with a Sepharose CL-4B (GE Healthcare) column. It is estimated that the PS was linked with about 30 Cy-5-Streptavidin molecules.

1.2. **Comparing Cy5-SA and Cy5-SA-PS conjugates**

The enhanced assay sensitivity using Cy5-SA-PS was tested by measuring TAb signal of SARS-CoV-2 negative serum spiked with monoclonal SARS-CoV-2 IgG (40150-D001, SinoBiological Inc., Wayne, PA) or IgM (Ab01680-15, Absolute Antibody, Boston, MA) at different concentrations (2.5-10 µg/mL). Cy5-SA conjugate (10 µg/mL), with or without addition of PS, is used as the signaling element. Experiments included 4 and 2 replicates for negative and spiked samples, respectively.

There are several aspects to the Cy5-SA-PS conjugate that enable enhanced assay sensitivity. Labeling the fluorescent dye initially to the streptavidin allows for control of the degree of substitution thereby avoiding over-labeling, which can quench the fluorescence signal. The PS, with its molecular weight about 5 million Daltons, serves as carrier of multiple Cy5-SAs (~ 30 SAs / PS), increasing the fluorescence signal when bound to the probe surface. SA is known to have a very high affinity for biotin. Therefore, since the PS carries multiple SAs, the avidity is increased further by binding to biotin on the probe surface. The PS is hydrophilic, which accounts for the very low background signal. Minimal non-specific binding to the probe surface is a critical feature in selecting the optimal high molecular weight carrier.

1.3. **The plaque reduction neutralization test (PRNT)**

Briefly, 100 µl of 2-fold serially diluted test sera were mixed with 100 µl of 200 plaque forming units (PFUs) of SARS-CoV-2, isolate USA-WA1/2020 (BEI Resources, NR181 52281) and incubated at 37°C in 5% CO_2_ for 1 h. Vero E6 cells (C1008, ATCC CRL-1586) were inoculated with the virus:serum mixture (100 ul) and adsorption was allowed to proceed for 1 h at 37ºC in in 5% CO_2_. A 0.6% agar overlay prepared in cell culture maintenance medium (Eagle’s Minimal Essential Medium, 2% heat-inactivated fetal bovine serum, 100 µg/ml Penicillin G, 100 U/ml Streptomycin) was added at the conclusion of adsorption A second agar overlay containing 0.2% Neutral red was added 2 days post-infection. After incubation for an additional one day, the number of plaques in each sample were recorded. The titer was reported as the inverse of the highest dilutions of sera providing 50% (PRNT50) reduction in the number of viral plaques, relative to virus-only infection.

1.4. **The pseudovirus neutralization test (PsV)**

To prepare SARS-CoV-2 pseudovirus stocks, 293T cells (American Tissue Culture Collection, ATCC) were seeded 24 h prior to transfection in a 150 cm^2^ flask at 60% confluency. An 18-µg aliquot of the pHIV-1_NL_ ΔEnv-NanoLuc plasmid (obtained from Paul Bieniasz, The Rockefeller University, New York) was co-transfected with 6 µg of pSARS-CoV-2-S_Δ19_ (which lacks the 19 C-terminal codons; also from Paul Bieniasz) using the Effectene (Qiagen) reagent^1^ . At 16 h after transfection, the cells were carefully washed twice and the medium replaced. Two days later, the cell supernatant (30 ml) was centrifuged at 1600 revolutions per minute for 10 min. The supernatant was then layered on to a 20% sucrose cushion and centrifuged at 200,000 G for 1.5 h at 4 ºC. The pseudovirus pellet was re-suspended in 30 ml of assay medium (Dulbecco’s Minimal Essential Medium supplemented with 10% fetal bovine serum, 20 IU/ml of Penicillin/Streptomycin (Gibco) and Glutamax (Gibco), and then aliquoted and frozen at -80 °C.

To conduct the assay, 96-well black tissue culture plates were coated with 50 µg/ml of Poly-L-Lysine for at least 2 h on the day before infection, then seeded with 1 x 10^4^ 293T/ACE2cl.22 cells/well (also from Paul Bieniasz). On the day of infection, serum or plasma samples (heat-inactivated earlier for 30 min at 56 ºC) were centrifuged using a microcentrifuge set at maximum speed for 10 min to remove particulate matter. The clarified samples were diluted 10-fold in assay medium and sterile filtered (Spin-X columns, Costar). They were then serially diluted in six 4-fold steps, mixed with pseudovirus, incubated at 37 ºC for 1 h and added to the target cells. The pseudovirus stocks had been previously titrated on the same cells. The amount added to the neutralization assay yielded a luciferase signal of 2 x 10^6^ relative light units (RLU) in the absence of serum or plasma. Each assay plate included virus control wells (cells and pseudovirus, no serum/plasma sample), viral input wells (pseudovirus only, no cells or sample) and cell-background wells (cells only, no pseudovirus or sample). Two days post-infection, the medium was carefully removed from each well before addition of Cell Lysis Buffer (Promega) at 50 µl/well. The plates were incubated for 15 min at room temperature on a shaker, NanoGlo substrate (Promega) was added (25 µl/well) and the plates were incubated for 5 min at room temperature. Luminescent counts per second (RLU) were detected using an Enspire reader (Perkin Elmer). The average RLU values derived from duplicate viral input wells were subtracted from duplicate values obtained using serum/plasma samples. The background-corrected values were plotted as percentage inhibition. The 8 virus control wells were used to determine the 100% infection level for each plate**.**

1.5. **Statistical analysis**

Bivariate associations between outcome variables (in-hospital mortality and intubation) and clinical parameters were evaluated using Fisher’s exact test or chi-squared test as appropriate between two categorical variables; t-test or Wilcoxon rank-sum test, as appropriate, between numerical variables. Correlations between two numerical variables were assessed by the Spearman correlation coefficient. Adjusted hazard ratios (HR, 95% confidence intervals) for in-hospital mortality and p-values were generated from Cox Proportional Hazards regression, adjusting for age and cancer comorbidity. Data are presented as mean ± SD or median with interquartile range (IQR) unless otherwise specified. p-values < 0.05 were considered significant. Analyses were performed in statistical programming language R version 4.0.2 (2020-06-22) or in GraphPad Prism Version 8.4.1 (GraphPad Software, La Jolla, CA).

2. **Supplemental Figures**

**Supplementary Figure 1. Linearity of (A) TOP TAb and (B) TOP SNAb.**

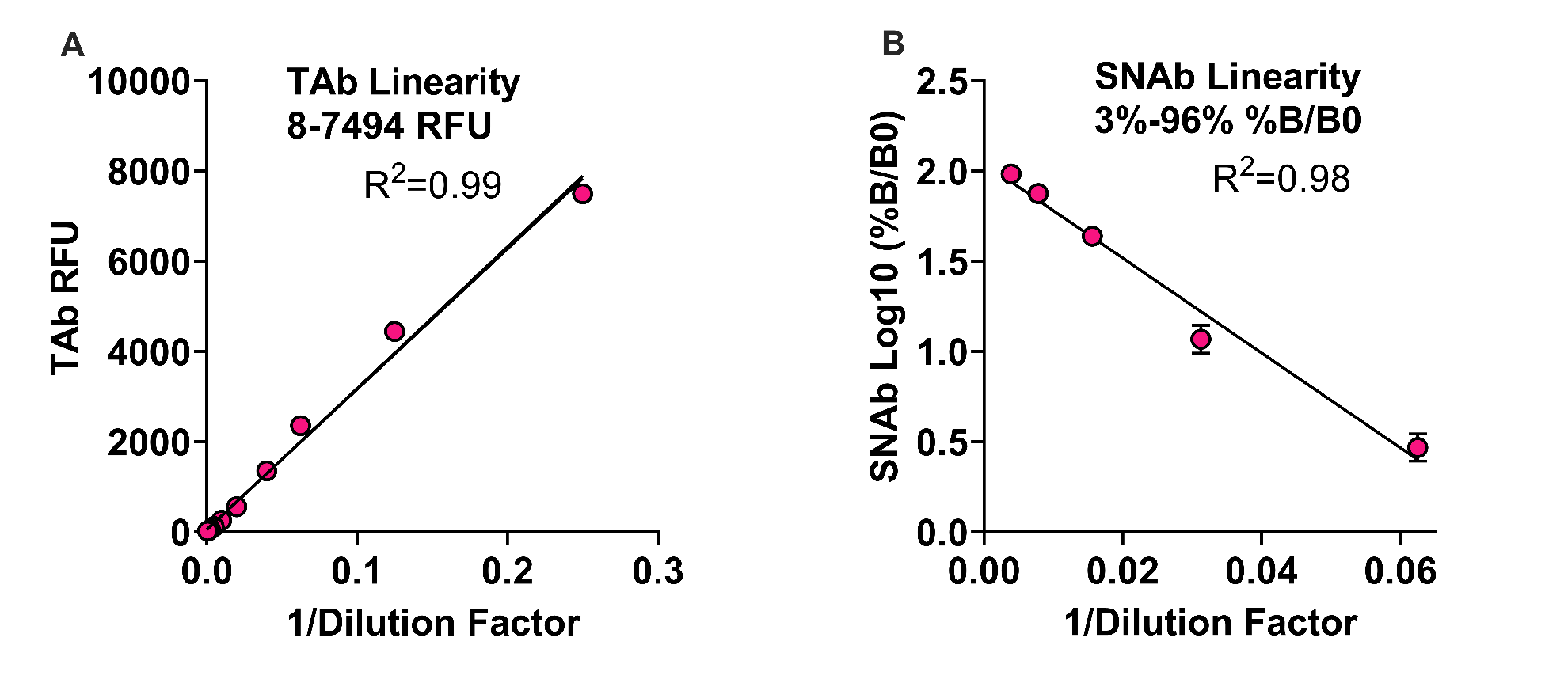

**Supplementary Figure 2. TOP-TAb and SNAb at initial ED presentation stratified by in-hospital intubation.** (A) TAb positive rate, (B) TAb level and (C) SNAb level in patients who were intubated or not intubated. No significant difference in antibody positivity rates or levels of TAb and SNAb in the initial ED samples was observed

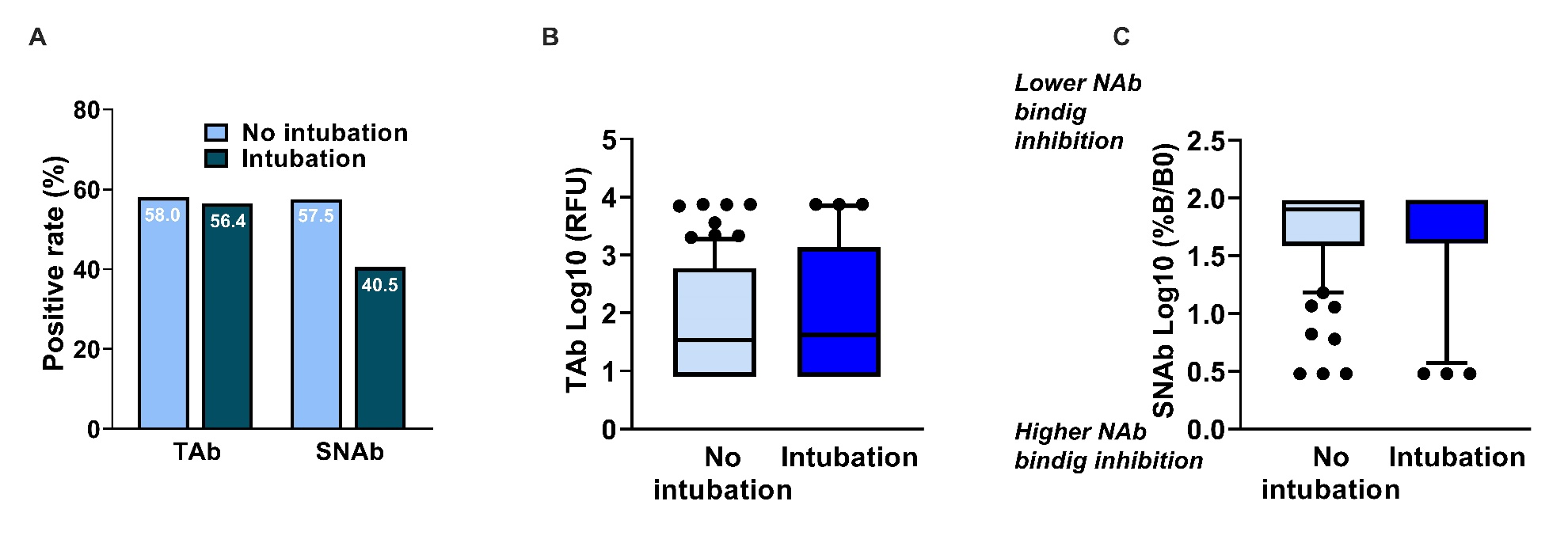

**3. Supplemental Tables**

**Supplemental Table 1. Detection rates of SARS-Cov-2 TOP-TAb, TOP-SNAb, and Roche TAb based on days after initial ED presentation.**

|  | **TOP-TAb** | **TOP-SNAb** | **Roche TAb** |
| --- | --- | --- | --- |
| Days after initial ED presentation: 0 | | | |
| Negative | 15 | 16 | 26 |
| Positive | 24 | 19 | 13 |
| Total | 39 | 35^*^ | 39 |
| Sensitivity % | 61.5 | 54.3 | 33.3 |
| 95% CI | 45.9-75.1 | 38.2-69.5 | 20.6-49.1 |
| p value (vs Roche TAb) | 0.023 | 0.100 |  |
| Days after initial ED presentation: 1-3 | | | |
| Negative | 5 | 9 | 11 |
| Positive | 33 | 27 | 25 |
| Total | 38 | 36^*^ | 36^*^ |
| Sensitivity % | 86.8 | 75.0 | 69.4 |
| 95% CI | 72.2-94.7 | 58.7-86.4 | 53.0-82.1 |
| p value (vs DAOS: 0) | 0.018 | 0.085 | 0.003 |
| p value (vs Roche TAb) | 0.092 | 0.793 |  |
| Days after initial ED presentation: 4-6 | | | |
| Negative | 5 | 4 | 8 |
| Positive | 34 | 33 | 30 |
| Total | 39 | 37^*^ | 38^*^ |
| Sensitivity % | 87.2 | 89.2 | 78.9 |
| 95% CI | 72.8-94.9 | 74.7-96.3 | 63.4-89.2 |
| p value (vs DAOS: 0) | 0.018 | 0.001 | <0.0001 |
| p value (vs Roche TAb) | 0.377 | 0.346 |  |

^*^Missing data was due to quantity not sufficient (QNS).

**Supplementary Table 2. Detection rates of SARS-Cov-2 TOP-TAb, TOP-SNAb, and Roche TAb based on days after onset of symptoms.**

|  | **TOP-TAb** | **TOP-SNAb** | **Roche TAb** |
| --- | --- | --- | --- |
| Days after onset of symptoms: 0-7 | | | |
| Negative | 15 | 19 | 25 |
| Positive | 20 | 12 | 9 |
| Total | 35 | 31^*^ | 34^*^ |
| Sensitivity % | 57.1 | 38.7 | 26.5 |
| 95% CI | 40.8-72.0 | 23.7-56.2 | 14.4-43.3 |
| p value (vs Roche TAb) | 0.015 | 0.426 |  |
| Days after onset of symptoms: 8-14 | | | |
| Negative | 9 | 9 | 17 |
| Positive | 50 | 47 | 42 |
| Total | 59 | 56^*^ | 59 |
| Sensitivity % | 84.7 | 83.9 | 71.2 |
| 95% CI | 73.3-92.0 | 72.0-91.5 | 58.5-81.2 |
| p value (vs DAOS: 0-7) | 0.006 | <0.0001 | <0.0001 |
| p value (vs Roche TAb) | 0.119 | 0.122 |  |
| Days after onset of symptoms: >14 | | | |
| Negative | 1 | 1 | 3 |
| Positive | 21 | 20 | 17 |
| Total | 22 | 21^*^ | 20^*^ |
| Sensitivity % | 95.5 | 95.2 | 85.0 |
| 95% CI | 76.5-100.0 | 75.6-100.0 | 63.1-95.6 |
| p value (vs DAOS: 0-7) | 0.002 | <0.0001 | <0.0001 |
| p value (vs Roche TAb) | 0.333 | 0.343 |  |

^*^Missing data was due to quantity not sufficient (QNS).

**Supplementary Table 3. Baseline characteristics of hospitalized patients stratified by intubation**

| **Variable** | **Categories/Statistics** | **Intubation**  **Yes (n=39)** | **Intubation**  **No (n=82)** | **P-value** |
| --- | --- | --- | --- | --- |
| Death | n. (%) | 21 (53.8%) | 12 (14.6%) | <0.001^C^ |
| Gender | Female- n. (%) | 8 (20.5%) | 34 (41.5%) | 0.024^C^ |
|  | Male- n. (%) | 31 (79.5%) | 48 (58.5%) |  |
| Age | Mean (SD) | 64.8 (14) | 66 (16.5) | 0.69^T^ |
|  | Median (IQR) | 68 (53.5, 72.5) | 67.5 (55.2, 78) |  |
|  | Range | (24, 90) | (24, 96) |  |
| Race | Asian- n. (%) | 5 (12.8%) | 2 (2.4%) | 0.39^F^ |
|  | Black/AA- n. (%) | 5 (12.8%) | 10 (12.2%) |  |
|  | Declined- n. (%) | 9 (23.1%) | 17 (20.7%) |  |
|  | Hispanic- n. (%) | 0 (0%) | 1 (1.2%) |  |
|  | Other- n. (%) | 5 (12.8%) | 15 (18.3%) |  |
|  | Unknown- n. (%) | 4 (10.3%) | 7 (8.5%) |  |
|  | White- n. (%) | 11 (28.2%) | 30 (36.6%) |  |
| DAOS at initial ED presentation | Mean (SD) | 8.5 (5.2) | 7.1 (4.5) | 0.26^E^ |
|  | Median (IQR) | 7 (4, 14) | 7 (3, 10) |  |
|  | Range | (1, 21) | (0, 20) |  |
| Cancer | n. (%) | 6 (15.4%) | 15 (18.3%) | 0.69^C^ |
| Other comorbidities^*^ | n. (%) | 27 (69.2%) | 61 (74.4%) | 0.55^C^ |
| Compromised immune status ^#^ | n. (%) | 4 (10.3%) | 10 (12.2%) | >0.99^F^ |
| Abdominal pain | n. (%) | 1 (2.6%) | 7 (8.5%) | 0.43^F^ |
| Fever | n. (%) | 21 (53.8%) | 56 (68.3%) | 0.12^C^ |
| Chest pain | n. (%) | 0 (0%) | 2 (2.4%) | >0.99^F^ |
| Shortness of breath | n. (%) | 27 (69.2%) | 45 (54.9%) | 0.13^C^ |
| Cough | n. (%) | 20 (51.3%) | 40 (48.8%) | 0.8^C^ |
| Diarrhea | n. (%) | 5 (12.8%) | 13 (15.9%) | 0.66^C^ |
| Headache | n. (%) | 2 (5.1%) | 4 (4.9%) | >0.99^F^ |
| Rhinitis | n. (%) | 0 (0%) | 2 (2.4%) | >0.99^F^ |
| Nausea/Vomiting | n. (%) | 6 (15.4%) | 10 (12.2%) | 0.63^C^ |
| Ageusia or Anosmia | n. (%) | 1 (2.6%) | 3 (3.7%) | >0.99^F^ |
| Body or muscle aches | n. (%) | 8 (20.5%) | 10 (12.2%) | 0.23^C^ |
| Fatigue/Weakness | n. (%) | 10 (25.6%) | 24 (29.3%) | 0.68^C^ |
| Sore throat | n. (%) | 2 (5.1%) | 0 (0%) | 0.1^F^ |

Test: C: Chi-square; E: Exact Wilcox; F: Fisher; T: T.test

^*^Other comorbidities included hypertension, type 2 diabetes, coronary artery disease, hyperlipidemia, and obesity

^#^ Compromised immune status included post-transplant, chemotherapy, radiation and chronic corticosteroid use

Abbreviations: SD: standard deviation; IQR: interquartile range, n: number; DAOS: days after onset of symptoms
